# Genetics of SSRI Antidepressant Use and Relationship to Psychiatric and Medical Traits

**DOI:** 10.1101/2025.03.20.25324264

**Authors:** Daniel Levey, Marco Galimberti, Joseph Deak, Priya Gupta, Stefany L. L. Empke, Keyrun Adhikari, Kelly Harrington, Rachel Quaden, J. J. Michael Gaziano, VA Million Veteran Program, Murray B. Stein, Joel Gelernter

## Abstract

Antidepressants are among the most-prescribed drugs worldwide, and selective serotonin reuptake inhibitors (SSRIs) are among the most prescribed antidepressants, most commonly used for major depression. We sought to increase our understanding of the biological relationships between SSRI use and a range of psychiatric traits by conducting Genome Wide Association Study (GWAS) in two large datasets, the UK Biobank (UKB) and the US Million Veteran Program (MVP). We conducted GWAS across 22 autosomes and the X chromosome in 777,952 individuals of European ancestry (191,800 SSRI users, 586,152 controls) and 112,526 individuals of African ancestry (53,499 SSRI users, 59,027 controls). We identified 40 genome-wide significant (GWS) loci, including two on the X chromosome. Using linkage disequilibrium score regression we detected strong correlations between MVP and the independent UKB cohort with use of specific SSRIs (fluoxetine genetic correlation (rg)=0.82 and citalopram rg=0.89) as well as with headaches (rg=0.80), major depressive disorder (MDD; rg=0.77), and spondylosis (rg=0.84), suggesting stability in the trait definition across cohorts. To evaluate differences between the genomic variance captured by the SSRI-use trait vs. MDD, we performed a comparative *rg* analysis between MDD and the meta-analysis for SSRI exposure and found significant differences, most notably for educational attainment (SSRI rg = -0.38, MDD rg= -0.26), cognitive performance (SSRI rg = -0.31, MDD rg=-0.15), and depression (SSRI rg = 0.80, MDD rg= 0.97). We compared locus discovery for SSRI use and MDD in the MVP, and found greater discovery for SSRI use (28 vs 17 risk loci). SSRI use is likely in part a proxy trait for MDD, while also presenting differences that may prove useful to disentangle MDD from other traits (e.g., anxiety disorders) that use similar pharmacological treatment.

## Introduction

Antidepressants are among the most-prescribed drugs worldwide [1], and selective serotonin reuptake inhibitors (SSRIs) are among the most prescribed antidepressants, commonly used for major depressive disorder (MDD), obsessive-compulsive disorder[2], anxiety disorders[3], and posttraumatic stress disorder[4], among other psychiatric disorders. They are widely prescribed by psychiatrists and primary care physicians, with variable response rates and tolerability.

While the antidepressant properties of the SSRIs are presumably related at least in part to inhibiting serotonin reuptake[5], there are many other possible mechanisms of action[6], and they have other biologically important properties as well. For example, many SSRIs are agonists at the sigma-1 receptor (S1R), a property which decreases inflammation via reducing endoplasmic reticulum stress.[7] This property identified SSRIs as compounds-of-interest for treatment of COVID19. Early in the COVID19 pandemic, drug repurposing was a major consideration, and numerous drugs were tested based on various theoretical considerations of in-vitro evidence, famously ivermectin and hydroxychloroquine, both of which failed rigorous clinical trials[8, 9] and appear to lack efficacy. Fluvoxamine, an SSRI that has S1R activity, was also evaluated, based on animal model evidence that S1R activity modulation could relate to inflammation and specifically that fluvoxamine reduces inflammatory response [10]. It was then tested in clinical samples, with initial success[11] and then replication in larger samples[12, 13] but inconsistent results for outpatient management[14, 15]. A subsequent study was negative[16]. Understanding the genetic relationship between SSRI use and COVID19 risk could clarify the biological picture further.

With the wide use of SSRIs, we can ask: what data are available readily, to increase our understanding of the biological relationships between SSRI use and a range of psychiatric traits? It would be possible to consider SSRI use *per se* in clinical samples, but we can also consider SSRI use as a genetic trait via querying the underlying genetic associations, including comparative genetic correlations with MDD.

What should we expect if we are to study the genetics of use of a certain pharmaceutical class of drugs? One possibility would be an attenuated set of findings directly relevant to the diagnostic class for which they are most frequently used; another would be a stronger set of findings for the diagnostic class because medication use might perhaps identify a more severe sub-phenotype. The phenotype could also reflect successful use of the medication, if those who are treated unsuccessfully discontinue use and are not designated with the “medication user” phenotype. Or the results could pertain to something more inherent to use of the class of medications as a meaningful genetic outcome, distinct from the diagnostic class or classes for which the medication is commonly used.

We set out to address the question of the genetic meaning of SSRI use both as a trait *per se* and how it fits with the biology and genetics of psychiatric traits, and with COVID19 traits, using individual data from the Million Veteran Program (MVP) and UK Biobank (UKB) samples of European (EUR) and African (AFR) ancestries and summary statistics from the COVID19 Host Genetics Initiative.

## Methods

### Phenotype

Tables containing de-identified participants with information on medications prescribed were acquired from the Veterans Affairs Electronic Health Record (EHR) to identify participants who had ever received SSRIs. We considered any SSRI prescription recorded in the inpatient or outpatient EHR for fluvoxamine, fluoxetine, sertraline, escitalopram, paroxetine, or citalopram to define a case for the primary analysis. After matching medication tables to participants with available genetic data and ancestry assignment, 177,494 cases (who had received SSRI prescriptions) and 268,353 controls (who had not) remained for analysis. In the UKB, cases were defined by use of SSRIs as described above from the treatment/medication code tables (data field 20003), and controls as subjects who had used none of these. The UKB data are from verbal interview on prescription medications by a trained nurse. After matching to unrelated EUR participants with genetic data there were 14,306 cases and 317,799 controls. We also explored AFR subjects within the MVP, of whom there were 53,499 cases and 59,027 controls.

### Genome-Wide Association Study (GWAS)

GWAS was performed by logistic regression using Plink2.0 software,[17] and has been described previously.[18, 19] Primary analyses considered use of any SSRI. Meta-analysis was conducted in the European Ancestry cohort using inverse variance weighting in METAL[20]. Sex stratified analysis of the X chromosome data was conducted in the MVP sample using XWAS 3.0[21] on hard call genotypes.

Following sex stratified analysis, test statistics from males and females were combined using Stouffer’s method[22].

### Linkage Disequilibrium Score Regression

Linkage Disequilibrium score regression (LDSC) was carried out using LDSC software[23]). Intercept and attenuation ratio were examined to test for inflation in GWAS results. Complex-Traits Genetic Virtual Lab (CTG-VL) [24] was used to investigate genetic correlations across 1,341 traits with GWAS summary statistics from MVP only (to avoid sample overlap with UKB) and tested into the range of traits available.

### Differential Correlation Analysis

To investigate genetic correlation (rG) differences between SSRI use and MDD to evaluate the relationship between these traits, LDSC analysis as above was repeated using summary statistics for a prior large study of MDD[25]. Differential rG was used to ascertain genetic differences between SSRI use and MDD, and was calculated by Z test for differences between the SSRI and the MDD meta-analyses[25] in the 1341 traits available in CTG-VL.

### Mendelian Randomization

We used MR to explore the relationship between EHR-identified ever-SSRI use and COVID19 related outcomes. Using data from the COVID Host Genome Consortium[26], we used leave-one-out summary statistics, omitting MVP data, for two trait definitions: 1. COVID19 infection vs population controls who had not reportedly been infected and 2. COVID19 infections resulting in hospitalization vs population controls who had not reportedly been infected. Leave-one-out (LOO) analysis removing the MVP cohort was only available for combined ancestry analysis as the LOO ancestry stratified analyses were only conducted removing 23andMe data, meaning 88%-89% of the cohort was EUR, depending on the phenotype. We also tested summary statistics from a GWAS of depression[25]. We used the R package TwoSampleMR to test four methods for MR analysis: MR-Egger, weighted median, inverse variance weighted (IVW), and simple mode. We used a p-value threshold of 5×10 for instrument selection, with default clumping window of 10 MB and clumping r2 cutoff of 0.001.

### Genomic Structural Equation Modeling

Genomic structural equation modeling (gSEM)[27] was used to perform EFA and CFA of SSRI use and 14 additional traits that were genetically correlated to examine further the meaning of the SSRI phenotype and to delineate similarities and differences with MDD. For EFA, factor structures composed of one to ten factors were examined. EFA model fit was evaluated by the amount of cumulative variance explained by the overall factor structure, the SS loadings (SS loading ≥1) for each included factor and the proportion of explained variance accounted for by each of the individual factors (that is, ≥10%). Traits with EFA factor loadings ≥0.20 were evaluated for optimal CFA model fit determined by conventional fit indices[27]. CFA models were estimated using diagonally weighted least squares estimation and a smoothed genetic covariance matrix. The 1KG phase 3 EUR reference panel was used for LD calculation.[28]

## Results

### GWAS

GWAS of the EUR ancestry cohorts for SSRI use resulted in 30 independent genome-wide significant (GWS) SNPs at 28 loci in MVP but no GWS signals in UKB, which had far fewer identified cases than MVP, 14,306 vs. 177,494. GWAS meta-analysis of the MVP and UKB cohorts yielded 40 total independent GWS SNPs (35 loci), including two loci on the X chromosome (Figure 1, Table 1). GWAS in the AFR MVP cohort yielded no GWS results.

**Figure 1.**
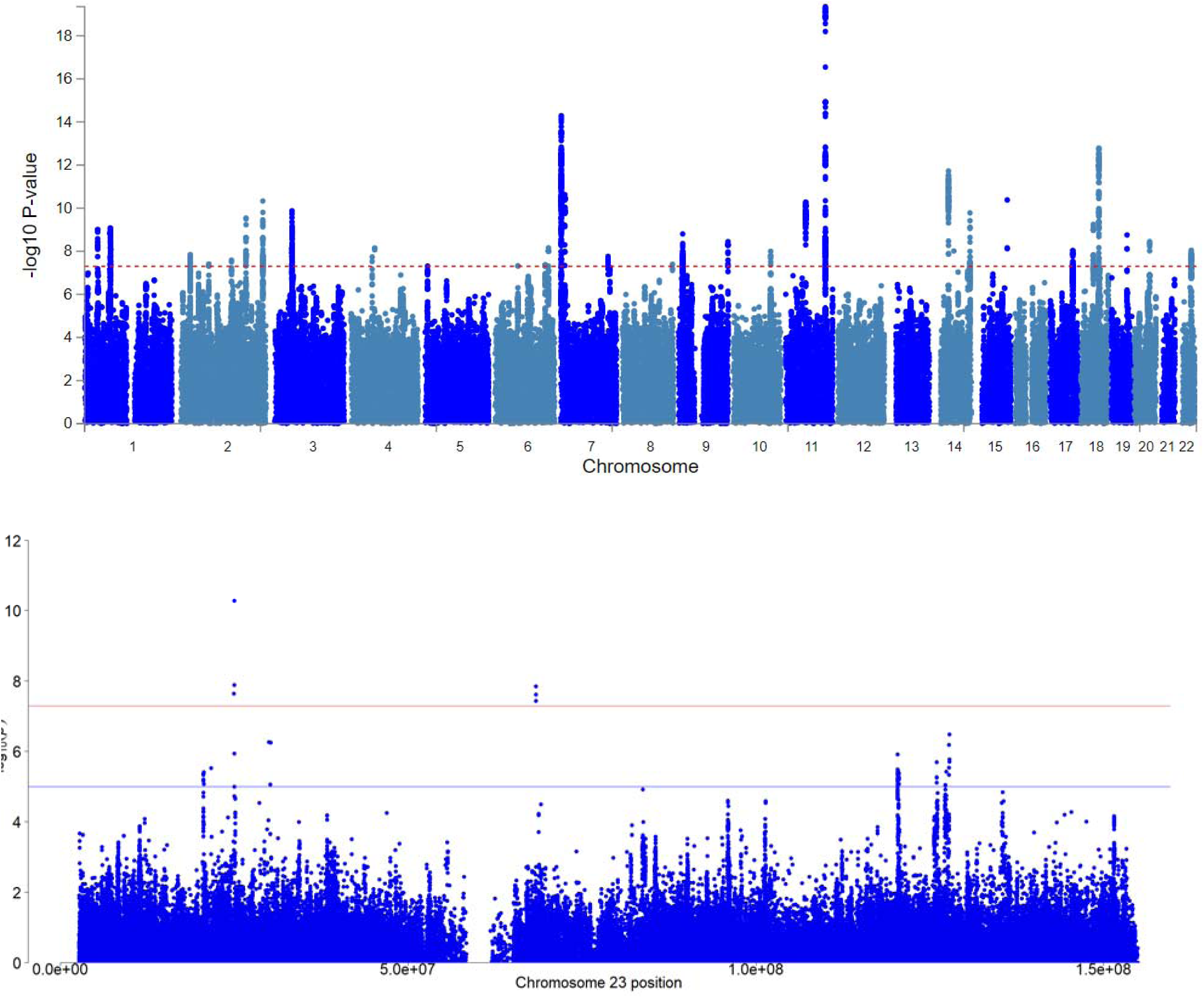
A. Manhattan plot for SSRI use in the autosomes. B. Focused plot for SSRI use on the X chromosome.

**Figure 2.**
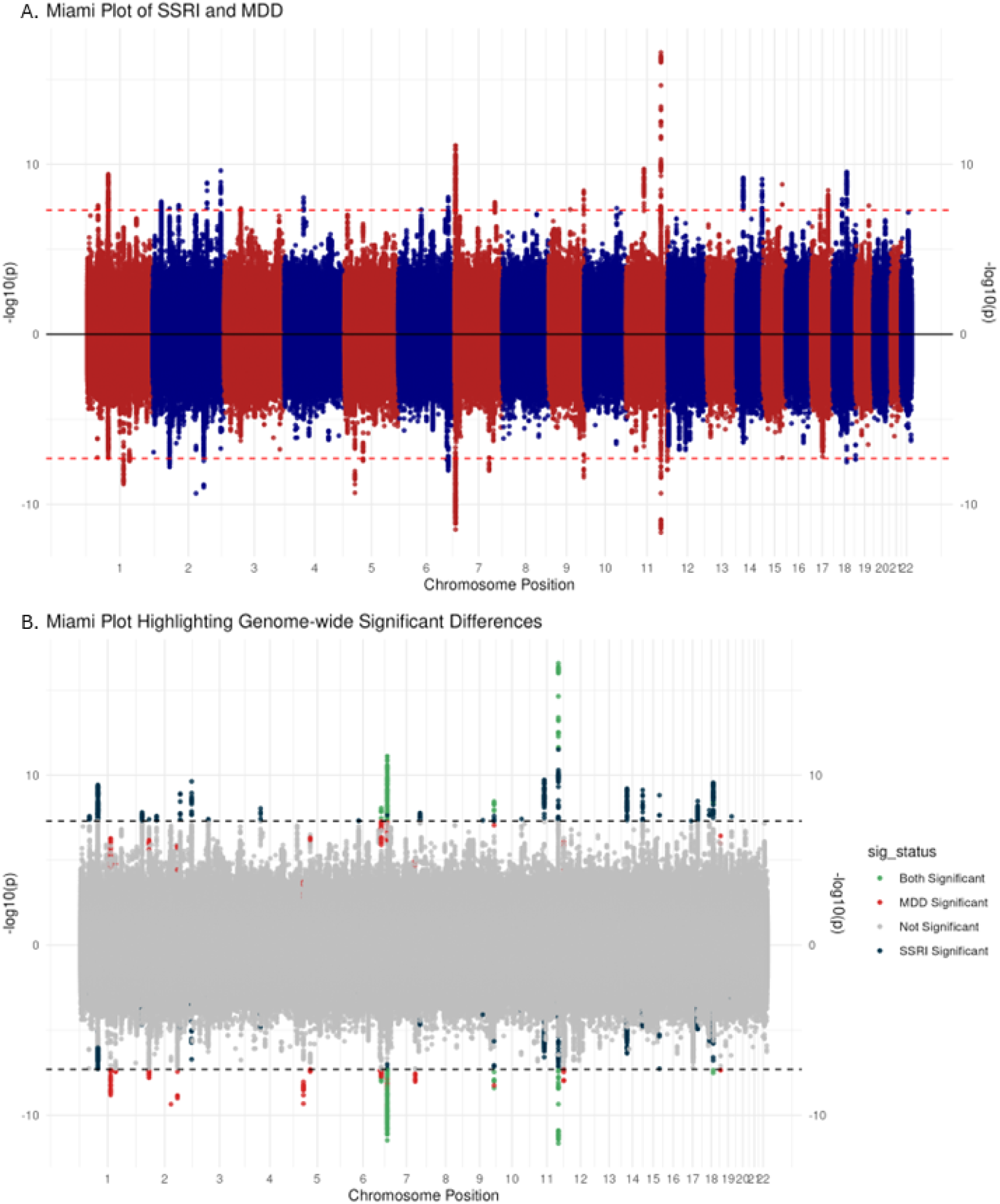
Comparing SSRI with MDD in the MVP. A. Miami plot depicts GWAS results for SSRI (above) and MDD (below). Both results are derived from the MVP cohort only. B. Miami plot highlighting differences identified, in the same cohort, between SSRI and MDD. GWS SNPs in both analyses are green, GWS SNPs from MDD are red, non significant SNPs in either are grey, and SNPs GWS in SSRI only are blue.

**Table 1.** Summary of GWAS results. 40 Independent loci depicted.

| No | rsID | chr | pos | p | Gene |
| --- | --- | --- | --- | --- | --- |
| 1 | rs4147222 | 1 | 37176422 | 9.94E-10 | GRIK3 |
| 2 | rs11209963 | 1 | 72883303 | 8.28E-10 | RPL31P12 |
| 3 | rs10890025 | 1 | 73738163 | 8.66E-10 | RN7SKP19 |
| 4 | rs7582361 | 2 | 27336376 | 1.43E-08 | CGREF1 |
| 5 | rs6707714 | 2 | 80007402 | 3.94E-08 | CTNNA2 |
| 6 | rs11677892 | 2 | 1.44E+08 | 2.58E-08 | ARHGAP15 |
| 7 | rs10931158 | 2 | 1.86E+08 | 2.87E-10 | ZNF804A |
| 8 | rs283466 | 2 | 2.34E+08 | 4.66E-11 | GIGYF2 |
| 9 | rs9586 | 3 | 49213637 | 1.33E-10 | KLHDC8B |
| 10 | rs143880703 | 4 | 59819356 | 9.12E-09 | RP11-506N2.1 |
| 11 | rs11725618 | 4 | 67053769 | 7.13E-09 | MIR1269A |
| 12 | rs113406393 | 5 | 7396327 | 4.91E-08 | ADCY2 |
| 13 | rs5876916 | 6 | 65433961 | 4.8E-08 | EYS |
| 14 | rs2077836 | 6 | 1.43E+08 | 4.31E-08 | RP11-440G9.1 |
| 15 | rs4869748 | 6 | 1.52E+08 | 6.96E-09 | ESR1 |
| 16 | rs11764590 | 7 | 2032803 | 5.2E-15 | MAD1L1 |
| 17 | rs13237518 | 7 | 12269593 | 2.35E-11 | TMEM106B |
| 18 | rs3812281 | 7 | 1.35E+08 | 1.75E-08 | CNOT4 |
| 19 | rs10098073 | 8 | 1.43E+08 | 4.06E-08 | TSNARE1 |
| 20 | rs17794133 | 9 | 11547243 | 1.58E-09 | PTPRD |
| 21 | rs11507683 | 9 | 1.4E+08 | 3.6E-09 | EXD3 |
| 22 | rs17078 | 10 | 1.07E+08 | 1.01E-08 | SORCS3 |
| 23 | rs10896661 | 11 | 57679029 | 5.2E-11 | CTNND1 |
| 24 | rs1940726 | 11 | 1.13E+08 | 2.25E-09 | NCAM1 |
| 25 | rs1554929 | 11 | 1.13E+08 | 1.45E-10 | DRD2 |
| 26 | rs34632468 | 11 | 1.13E+08 | 4.47E-20 | DRD2 |
| 27 | rs7125588 | 11 | 1.13E+08 | 8.71E-10 | DRD2 |
| 28 | rs67505447 | 14 | 42143231 | 1.89E-12 | LRFN5 |
| 29 | rs962961 | 14 | 57281154 | 9.71E-09 | OTX-AS1 |
| 30 | rs56101042 | 14 | 1.03E+08 | 1.65E-10 | TRAF3 |
| 31 | rs4702 | 15 | 91426560 | 4.18E-11 | FURIN |
| 32 | rs368956376 | 17 | 65906926 | 9.31E-09 | BPTF |
| 33 | rs7243428 | 18 | 35156177 | 5.66E-10 | CELF4 |
| 34 | rs71368910 | 18 | 50613671 | 1.97E-08 | DCC |
| 35 | rs72925341 | 18 | 50827476 | 1.65E-13 | DCC |
| 36 | rs6857 | 19 | 45392254 | 1.76E-09 | CTB-129P6.4 |
| 37 | rs9074 | 20 | 44688665 | 3.55E-09 | SLC12A5 |
| 38 | rs136402 | 22 | 41598933 | 8.98E-09 | L3MBTL2 |
| 39 | rs5944708 | X | 25000842 | 5.29E-11 | POLA1 |
| 40 | rs11539157 | X | 68381264 | 1.41E-08 | PJA1 |

**Table 2.** Mendelian randomization. Causal effect of SSRI exposure on COVID19 hospitalizations.

Hospitalized covid vs. population
| Exposure | Outcome | Method | nspns | b | se | pval |
| --- | --- | --- | --- | --- | --- | --- |
| SSRI | Covid | MR Egger | 124 | 0.04 | 0.13 | 0.77 |
| SSRI | Covid | Weighted median | 124 | 0.145 | 0.057 | 0.01 |
| SSRI | Covid | Inverse variance weighted | 124 | 0.204 | 0.039 | 1.4x10<sup>-7</sup> |
| SSRI | Covid | Simple mode | 124 | 0.44 | 0.17 | 0.009 |

### Linkage Disequilibrium Score Regression

SSRI use had significant SNP heritability (0.08 ± 0.004) as measured by LDSC. Genetic correlations calculated using LDSC showed highest rG with the MVP GWAS and UKB medication codes for two individual SSRIs, citalopram (rg=0.89) and fluoxetine (rg=0.82). These independent analyses, without sample overlap, indicate that the SSRI use trait is consistent across these populations. Notable significant positive correlations were also seen with headaches, informal detention (voluntary psychiatric inpatient), and MDD. The strongest negative associations were seen with for “no illness in siblings” as well as “not having bipolar or depression.” Significant negative correlation was also seen with age of mother’s death and age at first live birth. (Figure 3).

**Figure 3.**
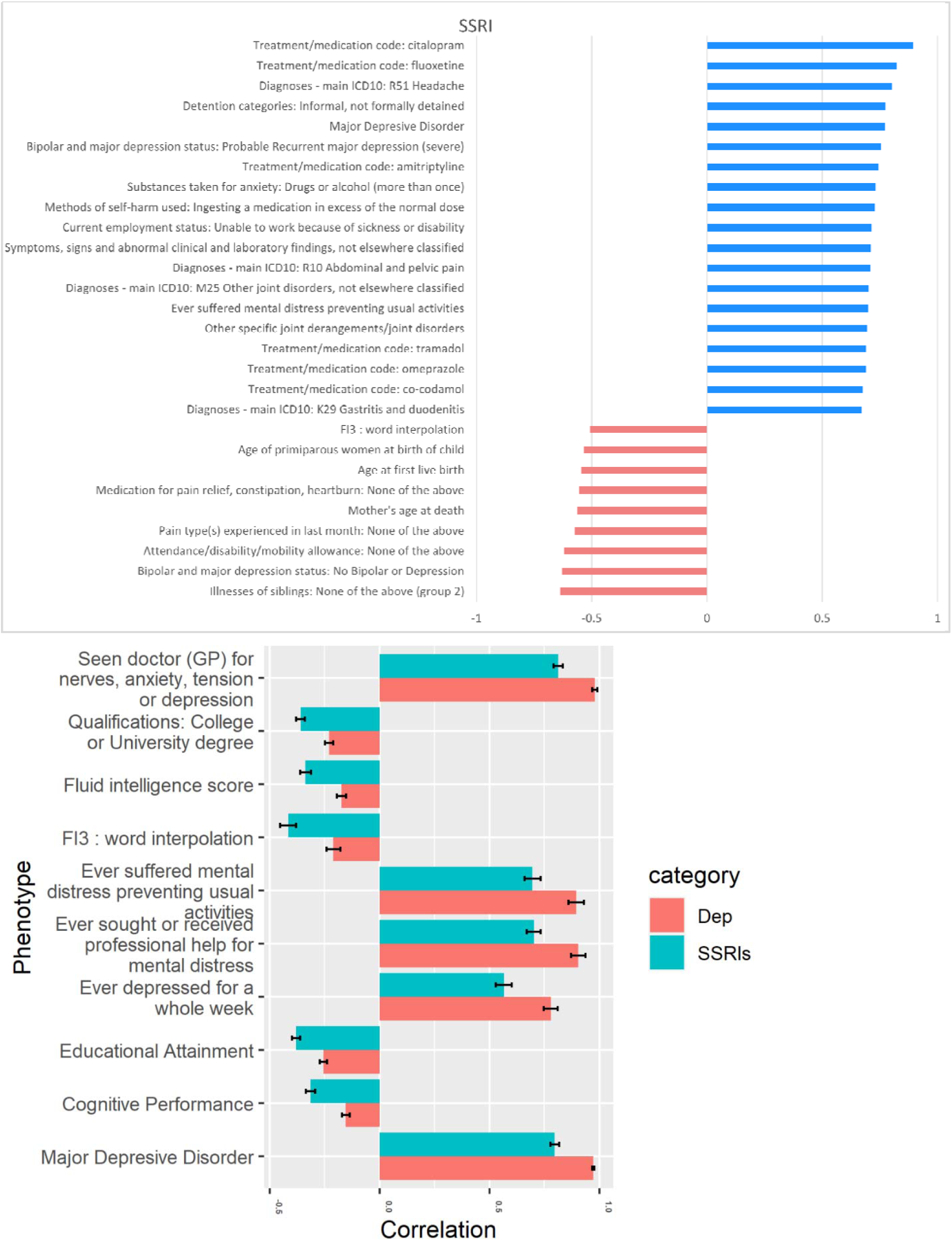
A Phenome-wide cross-trait rGs. B. Cross-trait Comparison of SSRI use trait to Depression.

### Differential Correlation Analysis

In the differential rG analysis comparing SSRI use and MDD, several traits showed different magnitudes of association, with nine of 1,314 tested remaining significant after multiple testing correction. Those traits were: “Major Depressive Disorder”, “seen doctor (GP) for nerves, anxiety, tension or depression”, “Cognitive Performance”, “Fluid intelligence score”, “Educational Attainment”, “Qualifications: College or University degree”, “Ever depressed for a whole week”, “Ever sought or received professional help for mental distress”, and “FI3 : word interpolation” (correct response as binary variable). (Figure 3).

### Mendelian Randomization

We investigated whether MDD could have a causal effect on COVID19 risk (considering the summary statistics from Hospitalized COVID19 or COVID19 infection vs population) and vice versa. Only the IVW method had a significant result (p=0.046) when considering the causality of MDD on COVID19 with a small positive effect (beta=0.083, se=0.042). All the other tests rejected the hypothesis (Table S3). We found significant positive causality with three of the four MR methods when testing the effect of SSRI use on COVID19 (Table S5), as can also be observed in the scatter and funnel plots (Figure S2-S3). We found no significant heterogeneity (pvalue MR Egger=0.20 and pvalue IVW=0.18 or pleiotropy (pvalue=0.17) from the sensitivity analyses. There was genetic correlation between SSRI use and COVID19 Hospitalizations (rg=0.21, se=0.05, pval=1.71×10^-5^). But no significant results were found when considering causal effects of COVID19 hospitalizations on SSRI use (Table S6). The summary statistics COVID19 infection vs population provided weaker results compared to Hospitalized COVID19 vs population (Table S3-S7) in MR analyses, but a similar genetic correlation with SSRI use (rg=0.27, se=0.05, pval=9.11×10^-8^). Both MDD and SSRI use were significant using the most powerful approach (IVW), while only SSRI use showed a significant effect with COVID19 for weighted median and simple mode sensitivity analyses. Neither were significant in the MR Egger analysis. MR analysis showed a bidirectional causal effect between SSRI and MDD (Table S1-S2).

### Genomic Structural Equation Modeling

From exploratory factor analysis (EFA), a four-factor model best fit the data, with the cumulative variance explained being 0.613, distributed relatively evenly across the four factors, with each accounting for between 12 % and 19% of the overall variance explained (factor 1 of 0.190, factor 2 of 0.182, factor 3 of 0.120 and factor 4 of 0.120). Each of the four factors had high sums of square (SS) loadings (factor 1 SS of 2.094, factor 2 SS of 2.007, factor 3 SS of 1.321 and factor 4 SS of 1.319). Factor 1 included PTSD symptoms (PCL-17) and the Generalized Anxiety Disorder 2-item symptom screener (GAD-2). Factor 2 featured suicide attempt, schizophrenia, and anorexia. Factor 3 contained BMI and COVID19, while factor 4 consisted of SSRI use. 3 traits co-loaded on factors, with cigarette smoking and Townsend Deprivation Index co-loading on factors 2 and 3 and MDD co-loading on factors 2 and 4 (Figure 4).

**Figure 4.**
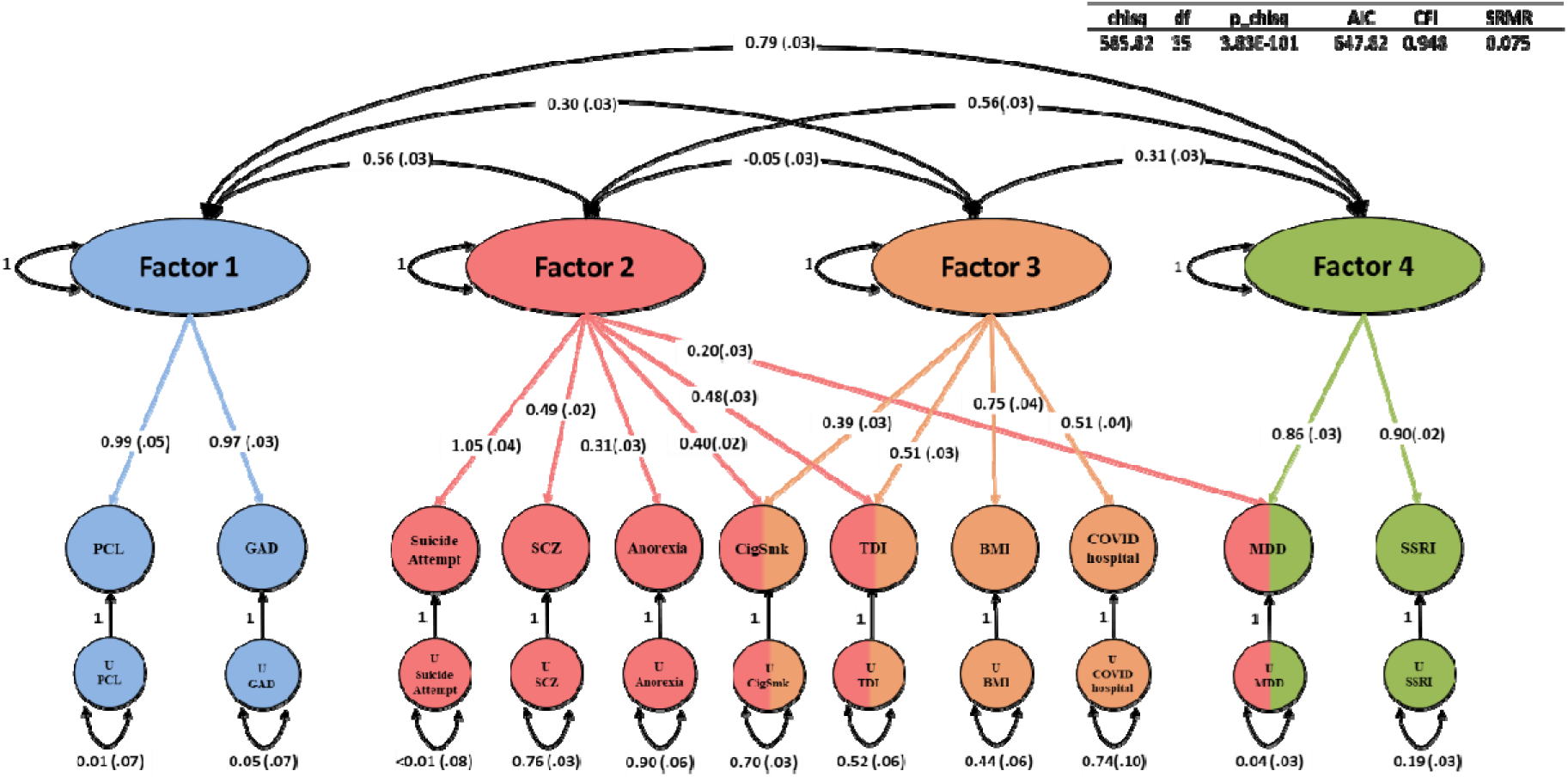
Genomic Structural Equation Model.

## Discussion

In this study, we approached the genetics and biology of psychiatric traits through studying SSRI use as the phenotype of interest. SSRI use provides a strong GWAS signal, with significant heritability.

Investigating the SSRI phenotype identified many more significant risk loci than investigating MDD (28 vs 17 in MVP EUR). SSRI use also had a nominally higher but similar SNP heritability (0.08 ± 0.004 vs 0.06 ± 0.003) than MDD. We were therefore sufficiently powered to be able to uncover underlying biology.

SSRI use in the MVP sample was highly genetically correlated with numerous other traits, but most strongly with use of specific SSRIs (citalopram and fluoxetine individually) in another sample, the UK Biobank. Strong correlations were observed despite differences between the samples (the MVP is lower SES and mostly male; the UKB is higher SES and includes more females than males.) Phenotype definition in the MVP was based on ever use of SSRI antidepressants while receiving care in the Veterans Affairs EHR, and in the UKB, was based on self-report of lifetime ever use.

Among the 40 independent significant SNPs identified, some are known from previous GWAS of related psychiatric traits. The most significant SNP in our GWAS was rs34632468, mapped to an intron of the gene encoding D2 dopamine receptor (*DRD2*). *DRD2* variants have been identified as risk loci for numerous substance use and psychiatric traits[29–31]; this SNP was identified recently in a large meta-analysis of tobacco use disorder.[32] The dopaminergic system features prominently in the neuroscience of addictive behaviors, and also with other psychiatric and behavioral phenotypes.[33, 34] A recent focus for MDD has been on perturbations in the pathways related to this reward system that may result in anhedonia, a core symptom of MDD.[35] Our lead finding in the gene encoding *FURIN*, rs4702, has been found at an intersection of many behavioral and psychiatry related traits such as insomnia[36], major depressive disorder[25], schizophrenia[37], opioid use disorder[38], and cannabis use disorder[39]. Work integrating GWAS and post-mortem eQTL expression data has highlighted a role for this specific SNP, and follow up analysis using CRISPR-mediated allelic conversion identified altered signaling in NGN2-excitatory neurons by multi-electrode array.[40]

Part of the reason for the under-reporting of data from X chromosome for many traits is due to the special considerations required to run the analysis accurately, relative to the autosomes. Unlike the autosomes, there is a sex difference in the number of copies of the chromosome: women have two copies while men only have one. This is common knowledge and basic biology, but it creates a problem for the assumptions of many common GWAS approaches. To account for this we used XWAS to conduct sex stratified analysis of the X chromosome and then combined by meta-analysis using Stouffer’s method. There were two X chromosome variant associations associated to SSRI use, identifying significant variation at rs5947708 in an intron of DNA polymerase alpha 1 catalytic subunit (*POLA1*), which plays an essential role in DNA replication, and at rs11539157 in exon 2 of the gene praja ring finger ubiquitin ligase 1 *(PJA1*), which has previously been linked to neurodevelopmental disorders [41] and to impaired learning and memory in mice.[42] Because both variants were located within the genes, we investigated further using the Combined Annotation Dependent Depletion database. Coding variant rs11539157 has a Phred-scaled score of 16.13, which is a rank ratio relative to other variants in the database, indicating this variant is in the top 4% of 9 billion potential SNVs in the database for deleterious effects. The X chromosome is historically understudied in GWAS analyses for technical reasons[43] despite containing genes of important function neurological function.[38] Continued work may identify that some of the missing heritability in GWAS studies may be from the practice of ignoring this chromosome.

Considered together, these GWAS and post-GWAS results are of interest regarding COVID19 risk and regarding the relationship of this trait to MDD. Understanding risk genes for SSRI use could also have value in evaluating medication response and pharmacogenomics. Efforts to bring personalized medicine to depression treatment have so far focused primarily on genetic variation in drug metabolizing enzymes – a panel including such variants was studied in the VA system and was shown to lead to statistically significant, but not necessarily clinically significant, improvement in outcome[44, 45]. Sustained SSRI use, as opposed to the binary trait we evaluated of use or no-use, could also be evaluated in future as a possible clinical predictor of treatment response.

We hypothesized that genetic instruments reflecting SSRI use might predict better COVID19 outcomes (as there is some prior evidence that fluvoxamine is efficacious for COVID19 infection), but we observed the opposite, that SSRI use (and MDD) predict worse outcomes. There are several possible explanations for this observation. One possible explanation is that SSRIs are not strongly effective for COVID19 treatment or prophylaxis – although it may protect from hospitalization. An additional explanation for the MR finding that SSRI use increases risk of COVID19 hospitalization is that we were indexing SSRI use over a long period, possible considering the long duration of the VA EHR – up to about three decades of use, in some cases comparable to lifetime measures. Data were collected prior to the outbreak of COVID19 and we have limited data post-COVID19 so we do not yet have a well-powered state-based temporal relationship to study. This is a trait, while the state (of current SSRI use) should be of greater relevance to SSRI-COVID19 risk interaction: that is, if SSRIs were truly effective for treating COVID19 infections, any effects on COVID19 risk or outcome should relate more closely to medication use at the time of COVID19 exposure; past use may not be relevant.

Another likely contributor to the positive association between SSRI use and COVID19 hospitalization is the high correlation (Rg=0.80) of SSRI use with MDD – MDD increases COVID19 risk[46]. Considering the high genetic correlation between MDD and SSRI use, SSRI use could be a proxy phenotype for MDD, although in this case it is hard to understand why the SSRI trait provided more locus discovery power than the depression trait. MR is often colloquially discussed as measuring causal effects of one trait on risk for another but more accurately, it measures the causal effect of the underlying genetic liability to a given trait. The strong “bidirectional” effect may indicate that SSRI and MDD are measuring related instruments from similar traits; genetic liability to one is similar to genetic liability to the other. When we compared GWAS for SSRI and MDD in the MVP cohort we observed many similarities but also interesting differences (Figure 2). While a locus on chromosome 5 is GWS only in MDD, a peak at the same position was observed to be just short of significance for SSRI use. It seems likely that this is a feature of statistical noise and random chance, and a larger follow up study might identify significant results for SSRI as well. Some differences may be more meaningful. We identified GWS variant associations on chromosomes 4, 14, 15 which showed no corresponding peaks in MDD, highlighting potential meaningful differences between these two related traits. The whole genome genetic correlation between the two traits as measured only within the MVP was 0.96. Future studies may consider the utility of drug prescriptions for an underlying condition as a useful proxy for the condition itself, but should also carefully consider the meaning and definition of the trait being studied.

Differential rG analysis between SSRI and MDD showed stronger negative rG with cognitive traits and fluid intelligence in the SSRI trait. Prior studies have shown greater MDD symptom severity is associated with reductions in cognitive function;[47] a possible explanation could be that this reflects that more severe symptoms are more likely to require that medication be dispensed.

Genetic correlation was highest with other SSRI use traits (specifically for citalopram and fluoxetine) and not with psychiatric traits typically *treated* by SSRIs. When this analysis was performed using MVP-only data the cross-cohort genetic overlap was substantially high with UKB traits for specific SSRIs (rGxCitalopram=0.90; rGxFluoxetine=0.82). While MDD in particular showed an rG with SSRI of 0.80, rG with citalopram was nearly 1 (rG=0.97). SSRI use is a heterogenous trait encompassing several underlying conditions, but even so, it seems remarkably consistent across cohorts.

Genomic structural equation modeling was used to understand the interrelations between associated complex traits better. Unsurprisingly, SSRI use clustered most strongly into a factor with MDD. This cluster was itself strongly correlated with an anxiogenic cluster of anxiety and post-traumatic stress disorder. Post-traumatic stress disorder was formerly considered to be an anxiety disorder in the previous iteration of the Diagnostic and Statistical Manual, DSM-IV[48] for diagnosing psychiatric disorders. Both anxiety disorders[49] and PTSD[50] are sometimes treated with SSRIs in the clinic.

MDD, but not SSRI, co-loads onto an additional factor that is composed of schizophrenia, suicide attempt, and anorexia. COVID19 hospitalization loads most strongly on a third factor with traits often associated with physical well-being such as BMI, the Townsend Deprivation Index, and smoking.

This study has a number of limitations. We assessed SSRI use, which is a complex trait that could have many causal trajectories that were not measured. We did not study the time course of treatment, proximal cause for SSRI prescription, number of prescriptions filled, treatment adherence, or other factors relating to the rationale for, and longitudinal course of, SSRI used. These extra dimensions could provide additional context and insight into the genetic associations identified for SSRI use. Additionally, “ever-use” of an SSRI (either lifetime or over the course of available EHR prescribing data) does not capture the essential temporal element that might be required for use as a proxy for possible SSRI effectiveness in the treatment of COVID19. The only available dataset for COVID19 phenotypes containing non-overlapping samples contained some non-EUR ancestry subjects integrated into the dataset, accounting for 11-12% of the total sample depending upon the phenotype and slightly limiting interpretation of these data.

In conclusion, we investigated SSRI use as a trait, using data from the MVP and the UK Biobank. We identified more loci for SSRI use than for MDD as well as nominally higher SNP heritability for the former trait, despite the apparent higher underlying phenotypic heterogeneity initially expected to characterize this trait – arguably SSRI use should encompass both the heterogeneity of MDD, plus that of other conditions for which SSRIs are prescribed. We included analysis of the X chromosome, a portion of the genome which remains understudied in genome-wide analyses, identifying two novel loci associated with SSRI use. We performed a comparative MR investigation between MDD and SSRI use with COVID19 infection and hospitalizations, identifying an effect of genetic liability of exposure to MDD and SSRI on both outcomes. In gSEM analysis SSRI use loaded most strongly onto a single factor while MDD co-loaded onto this same factor but also with a second factor associated with schizophrenia, anorexia and suicide attempt but not with anxiety or PTSD symptoms. Treatment-related traits such as SSRI use may be of substantial value in disentangling the genetic architecture of complex traits.

## Data Availability

Data will be made available from PI lab websites upon publication: https://medicine.yale.edu/lab/leveylab/data/
MVP specific data will be available through dbGAP: https://www.ncbi.nlm.nih.gov/projects/gap/cgi-bin/study.cgi?study_id=phs001672.v12.p1

## Tables/Figures

**Table S1:** Causal effect of SSRI on depression by MR. nsnps refers to the number of SNPs.

| Exposure | Outcome | Method | nsnps | B | se | pval |
| --- | --- | --- | --- | --- | --- | --- |
| SSRI | Depression | MR Egger | 121 | 0.642 | 0.078 | $2.09 \times 10^{-13}$ |
| SSRI | Depression | Weighted median | 121 | 0.720 | 0.038 | $4.00 \times 10^{-82}$ |
| SSRI | Depression | Inverse variance weighted | 121 | 0.767 | 0.023 | $3.45 \times 10^{-254}$ |
| SSRI | Depression | Simple mode | 121 | 0.69 | 0.11 | $1.76 \times 10^{-9}$ |

**Table S2:**
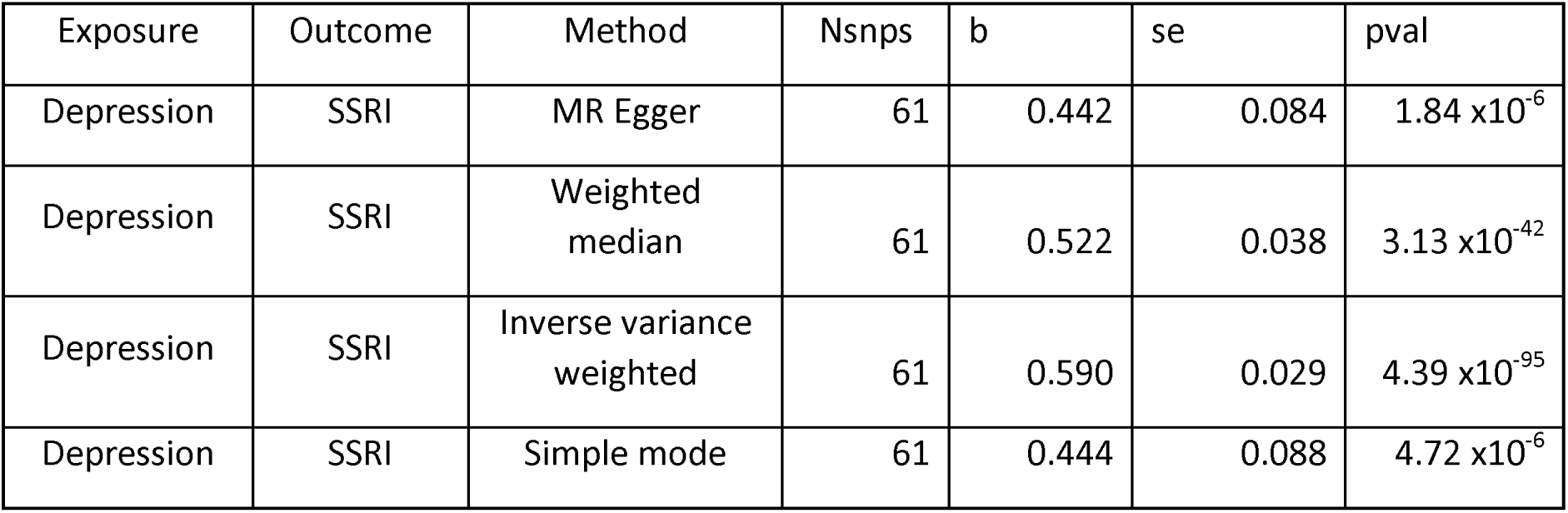
Causal effect of depression on SSRI by MR. nsnps refers to the number of SNPs.

**Table S3:**
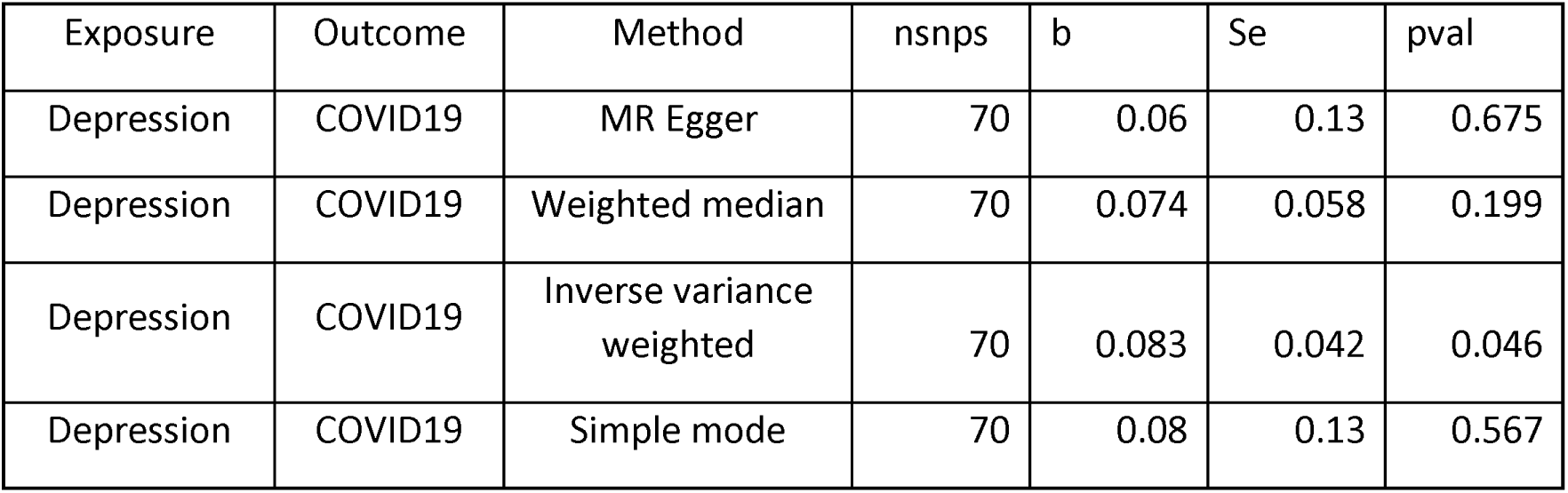
Causal effect of depression on COVID19 by MR. nsnps refers to the number of SNPs. For COVID19 it refers to the summary statistics *Hospitalized COVID19 vs population*.

**Table S4:**
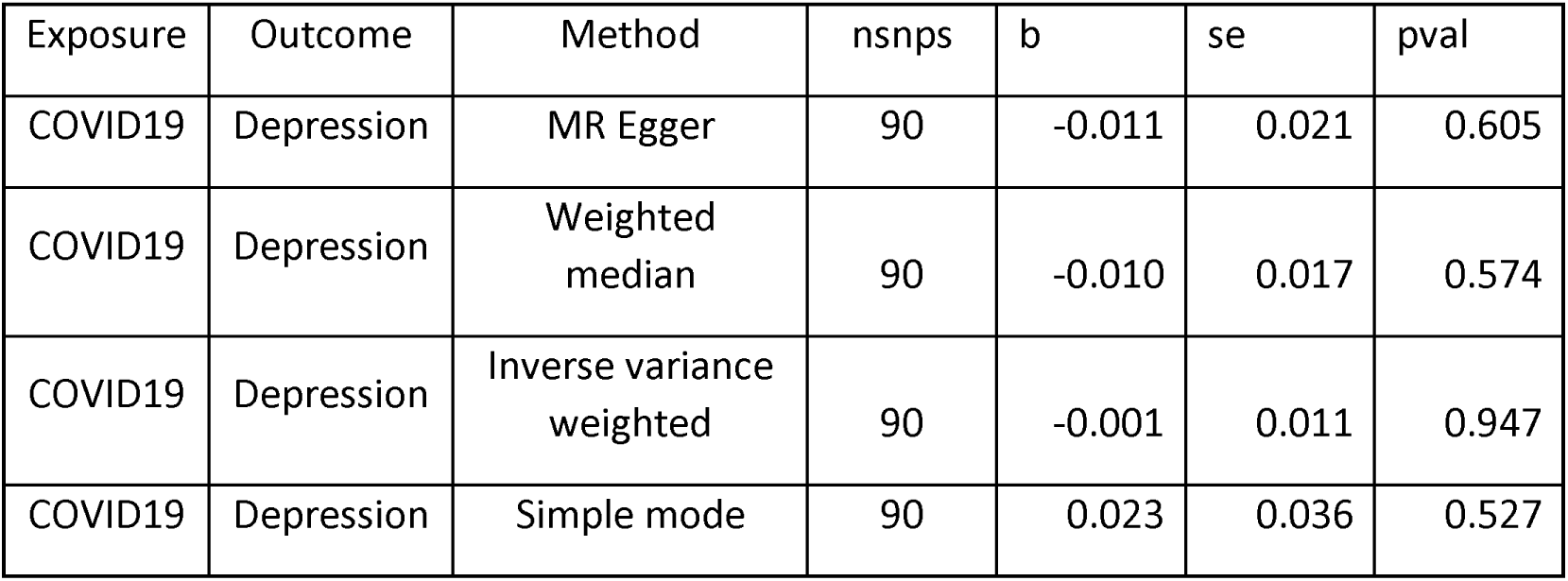
Causal effect of COVID19 on depression by MR. nsnps refers to the number of SNPs. For COVID19 it refers to the summary statistics *Hospitalized COVID19 vs population*.

**Table S5:**
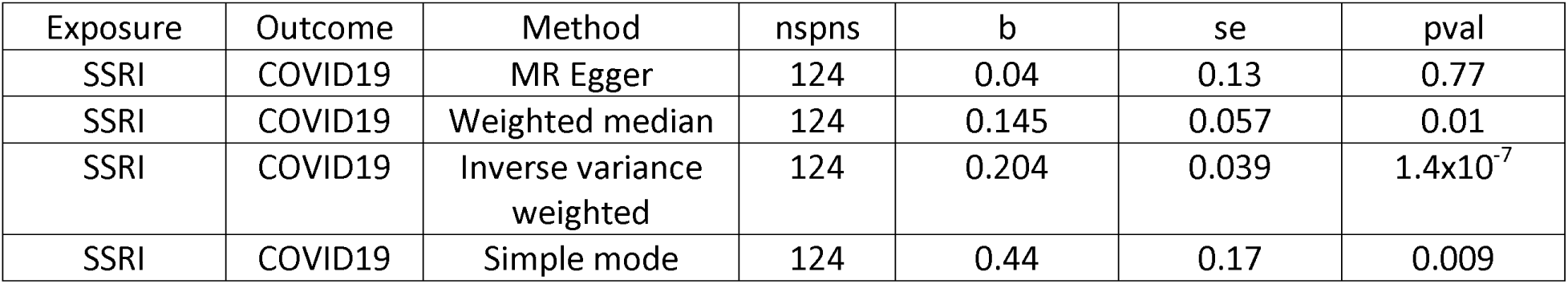
Causal effect of SSRI on COVID19 by MR. nsnps refers to the number of SNPs. For COVID19 it refers to the summary statistics *Hospitalized COVID19 vs population*.

**Table S6:**
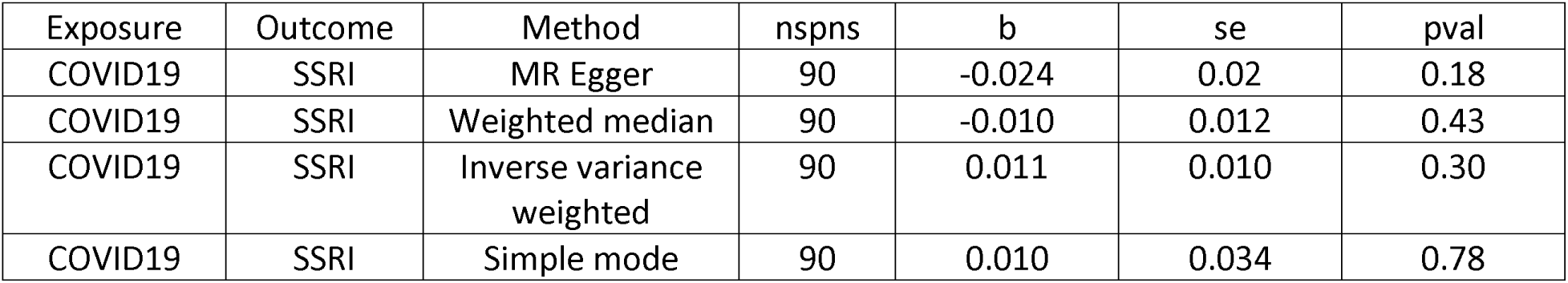
Causal effect of COVID19 on SSRI by MR. nsnps refers to the number of SNPs. For COVID19 it refers to the summary statistics *Hospitalized COVID19 vs population*.

**Table S7:**
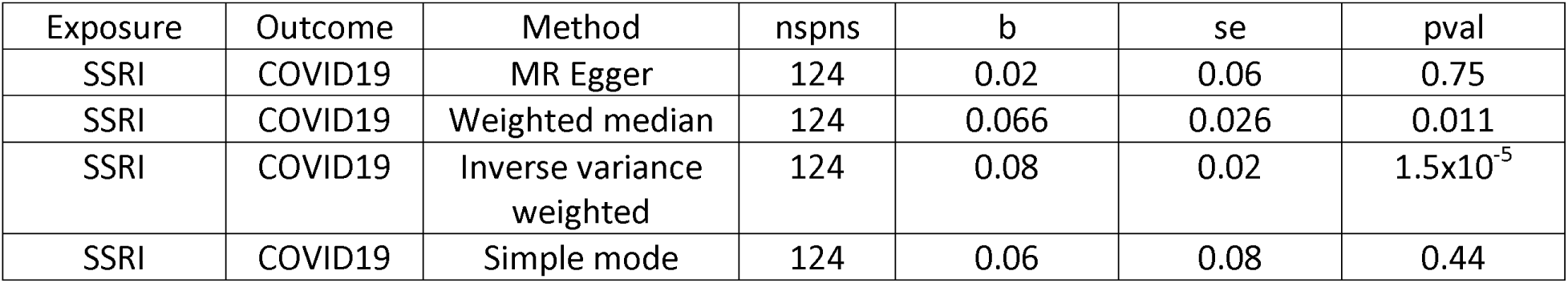
Causal effect of SSRI on COVID19 by MR. nsnps refers to the number of SNPs. For COVID19 it refers to the summary statistics *COVID vs population*.

**Table S8:**
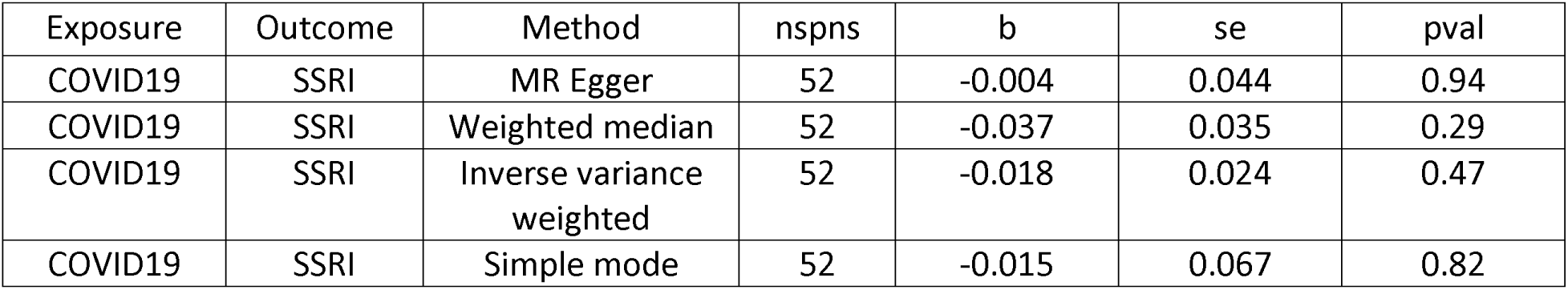
Causal effect of COVID19 on SSRI by MR. nsnps refers to the number of SNPs. For COVID19 it refers to the summary statistics *COVID vs population*.

**Figure S1:**
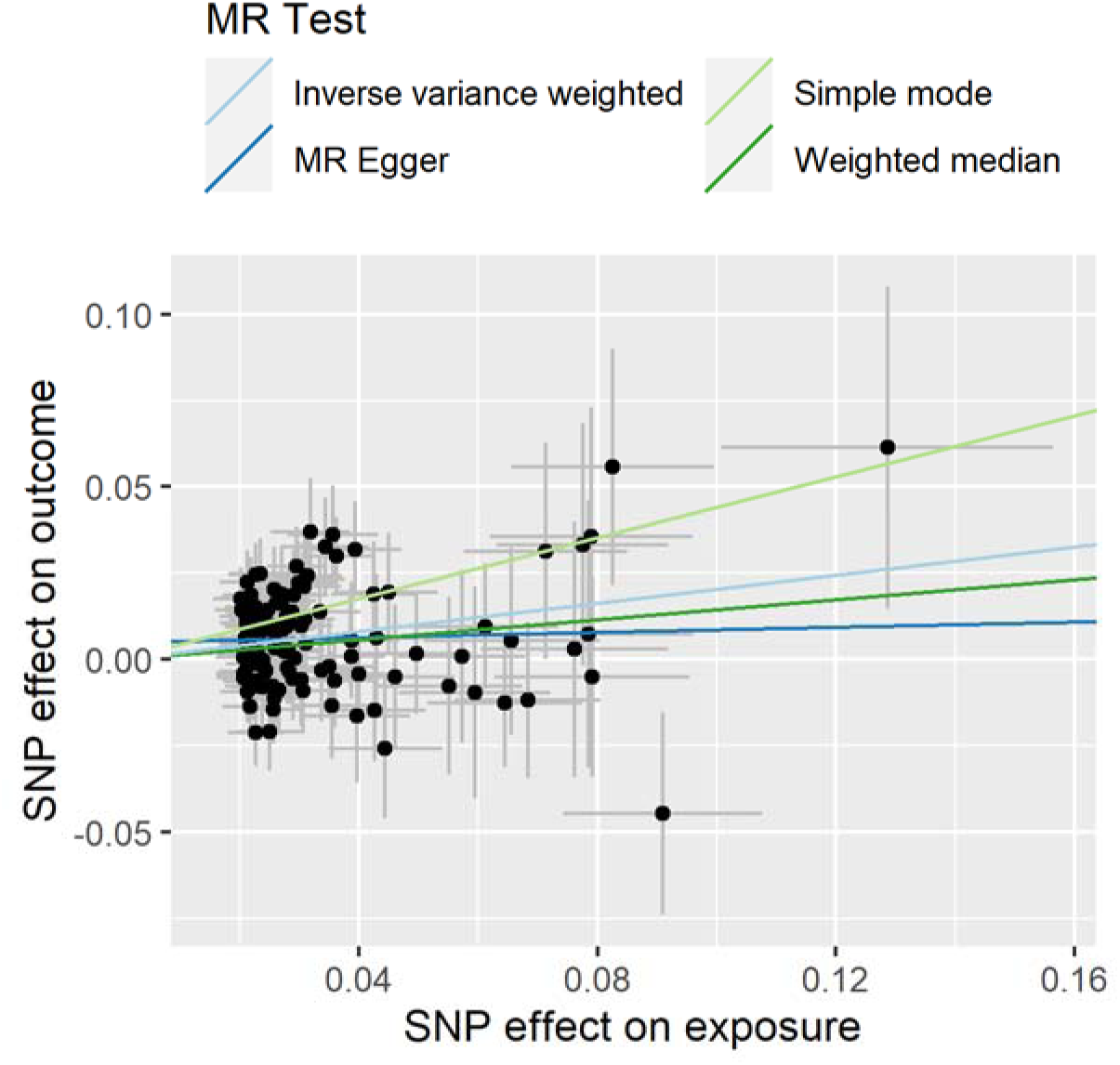
Scatter plot of SSRI as exposure and COVID19 as outcome. For COVID19 it refers to the summary statistics *Hospitalized COVID19 vs population*.

**Figure S2:**
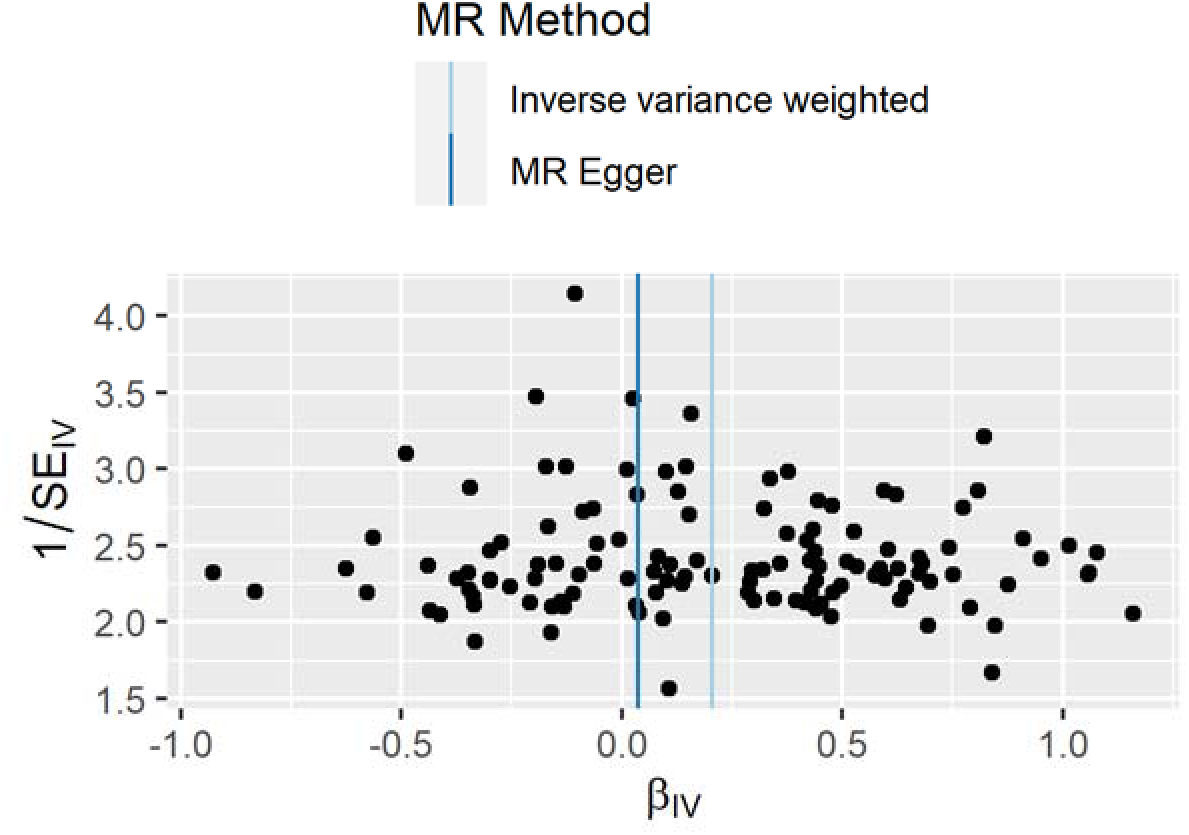
Funnel plot of SSRI as exposure and COVID19 as outcome. For COVID19 it refers to the summary statistics *Hospitalized COVID19 vs population*.

**Figure S3.**
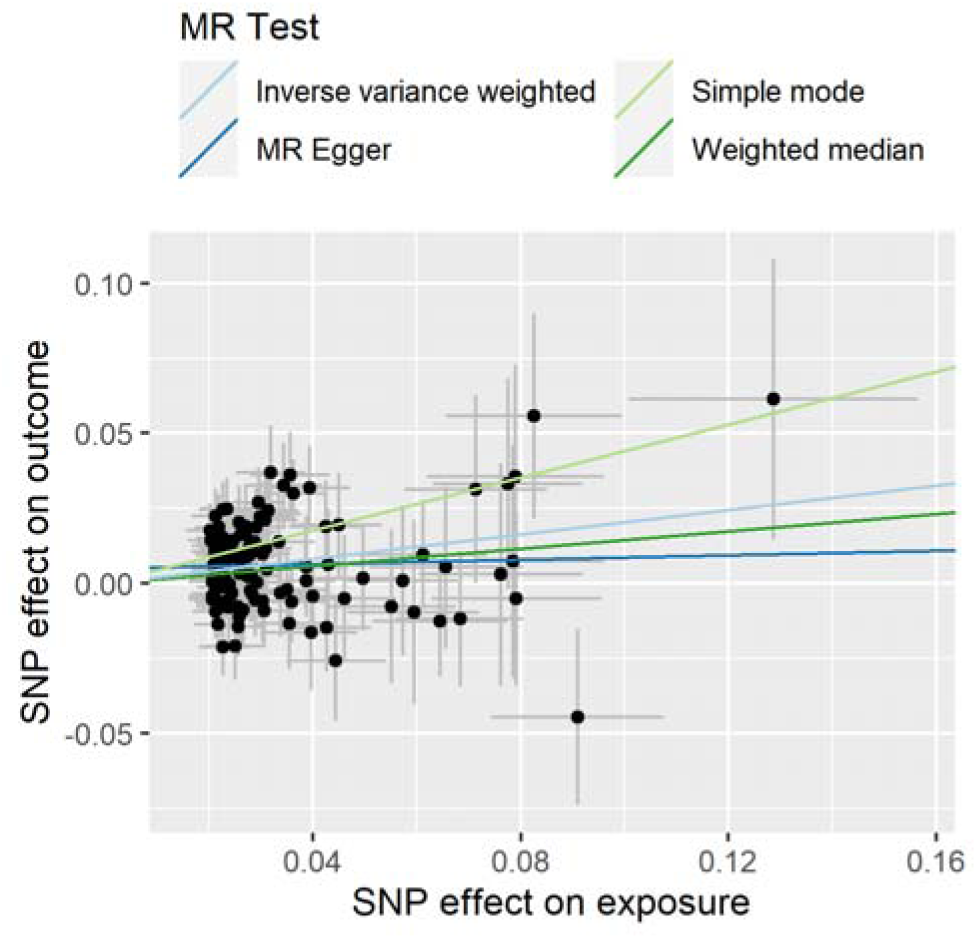
SNP effects from Mendelian randomization study of SSRI exposure on COVID19 hospitalization.

## Acknowledgements

This research is based on data from the Million Veteran Program, Office of Research and Development, Veterans Health Administration, and was supported by MVP092 and MVP069 as well as award #2IO1BX006482 and #5IK2BX005058. This publication does not represent the views of the Department of Veteran Affairs or the United States Government.

## VA Million Veteran Program

### Core Acknowledgements for Publications May 2024

#### MVP Program Office

- Sumitra Muralidhar, Ph.D., Program Director

US Department of Veterans Affairs, 810 Vermont Avenue NW, Washington, DC 20420

- Jennifer Moser, Ph.D., Associate Director, Scientific Programs

US Department of Veterans Affairs, 810 Vermont Avenue NW, Washington, DC 20420

- Jennifer E. Deen, B.S., Associate Director, Cohort & Public Relations

US Department of Veterans Affairs, 810 Vermont Avenue NW, Washington, DC 20420

#### MVP Executive Committee

- Co-Chair: Philip S. Tsao, Ph.D.

VA Palo Alto Health Care System, 3801 Miranda Avenue, Palo Alto, CA 94304

- Co-Chair: Sumitra Muralidhar, Ph.D.

US Department of Veterans Affairs, 810 Vermont Avenue NW, Washington, DC 20420

- J. Michael Gaziano, M.D., M.P.H.

VA Boston Healthcare System, 150 S. Huntington Avenue, Boston, MA 02130

- Elizabeth Hauser, Ph.D.

Durham VA Medical Center, 508 Fulton Street, Durham, NC 27705

- Amy Kilbourne, Ph.D., M.P.H.

VA HSR&D, 2215 Fuller Road, Ann Arbor, MI 48105

- Michael Matheny, M.D., M.S., M.P.H.

VA Tennessee Valley Healthcare System, 1310 24th Ave. South, Nashville, TN 37212

- Dave Oslin, M.D.

Philadelphia VA Medical Center, 3900 Woodland Avenue, Philadelphia, PA 19104

- Deepak Voora, MD

Durham VA Medical Center, 508 Fulton Street, Durham, NC 27705

#### MVP Co-Principal Investigators

- J. Michael Gaziano, M.D., M.P.H.

VA Boston Healthcare System, 150 S. Huntington Avenue, Boston, MA 02130

- Philip S. Tsao, Ph.D.

VA Palo Alto Health Care System, 3801 Miranda Avenue, Palo Alto, CA 94304

#### MVP Core Operations

- Jessica V. Brewer, M.P.H., Director, MVP Cohort Operations

VA Boston Healthcare System, 150 S. Huntington Avenue, Boston, MA 02130

- Mary T. Brophy M.D., M.P.H., Director, VA Central Biorepository

VA Boston Healthcare System, 150 S. Huntington Avenue, Boston, MA 02130

- Kelly Cho, M.P.H, Ph.D., Director, MVP Phenomics

VA Boston Healthcare System, 150 S. Huntington Avenue, Boston, MA 02130

- Lori Churby, B.S., Director, MVP Regulatory Affairs

VA Palo Alto Health Care System, 3801 Miranda Avenue, Palo Alto, CA 94304

- Scott L. DuVall, Ph.D., Director, VA Informatics and Computing Infrastructure (VINCI) VA Salt Lake City Health Care System, 500 Foothill Drive, Salt Lake City, UT 84148

- Saiju Pyarajan Ph.D., Director, Data and Computational Sciences

VA Boston Healthcare System, 150 S. Huntington Avenue, Boston, MA 02130

- Robert Ringer, Pharm.D., Director, VA Albuquerque Central Biorepository

New Mexico VA Health Care System, 1501 San Pedro Drive SE, Albuquerque, NM 87108

- Luis E. Selva, Ph.D., Director, MVP Biorepository Coordination

VA Boston Healthcare System, 150 S. Huntington Avenue, Boston, MA 02130

- Shahpoor (Alex) Shayan, M.S., Director, MVP PRE Informatics

VA Boston Healthcare System, 150 S. Huntington Avenue, Boston, MA 02130

- Brady Stephens, M.S., Principal Investigator, MVP Information Center Canandaigua VA Medical Center, 400 Fort Hill Avenue, Canandaigua, NY 14424

- Stacey B. Whitbourne, Ph.D., Director, MVP Cohort Development and Management VA Boston Healthcare System, 150 S. Huntington Avenue, Boston, MA 02130

#### MVP Publications and Presentations Committee

- Co-Chair: Themistocles L. Assimes, M.D., Ph. D

VA Palo Alto Health Care System, 3801 Miranda Avenue, Palo Alto, CA 94304

- Co-Chair: Adriana Hung, M.D.; M.P.H

VA Tennessee Valley Healthcare System, 1310 24 Ave. South, Nashville, TN 37212

- Co-Chair: Henry Kranzler, M.D.

Philadelphia VA Medical Center, 3900 Woodland Avenue, Philadelphia, PA 19104

